# Generative AI Versus Primary Care Physicians from Plan Initiation to Long-Term Maintenance of Chronic Disease Self-Management: A Vignette-Based Validation Study

**DOI:** 10.64898/2026.11.10.26367736

**Authors:** Ziyin Zhao, Huiming Yang, Qiting Wang, Ying Wang, Weibin Cheng, Xiaochen Ma

**Affiliations:** School of Public Health, Peking University, Beijing, China; China Center for Health Development Studies, Peking University, Beijing, China; National Health Commission Key Laboratory of Health System Reform and Governance, Peking University, Beijing, China; School of Public Health, Guangdong Pharmaceutical University, Guangdong, China; Institute for Healthcare Artificial Intelligence Application, Guangdong Second Provincial General Hospital, Guangdong, China; Department of Medical Statistics and Center for Health Information Research and Guangdong Key Laboratory of Medicine, School of Public Health, Sun Yat-Sen University, Guangdong, China

**Keywords:** generative artificial intelligence, chronic disease, self-management, validation study

## Abstract

**Background:** Generative artificial intelligence (GenAI) may provide personalised and dynamically adjustable support for chronic disease self-management. However, previous studies have largely focused on isolated questions and a limited range of evaluation dimensions.

**Objective:** To compare the quality of self-management advice generated by GenAI and primary care physicians (PCPs) across multiple stages of chronic disease management and evaluation dimensions.

**Methods:** We conducted a cross-sectional, source-blinded comparative evaluation using 15 standardised vignettes of diabetes and hypertension derived from authentic primary care follow-up records. Each vignette comprised three sequential management stages, with medical accuracy, readability, and behaviour-change potential assessed across the three simulated stages; individualisation and multidisciplinary integration at plan initiation; contextual adaptability during acute adjustment; and empathy during long-term maintenance. The GenAI system and 22 PCPs independently generated advice which was independently rated by three clinicians, one each from general practice, endocrinology, and cardiology. across seven dimensions. Linear mixed-effects models were used for the primary comparisons, and case-level paired and physician-distribution analyses were conducted as robustness analyses.

**Results:** The GenAI system achieved estimated marginal means above 4.0 in all dimensions except readability. It scored significantly higher than PCPs in six of the seven dimensions, with adjusted mean differences ranging from 1.16 to 1.56 points (P<.001). Readability was the only exception, with no significant difference between GenAI and PCPs (−0.22, 95% CI −0.54 to 0.11; P=.19). The GenAI mean exceeded the individual means of all recruited PCPs in the six dimensions but ranked at approximately the 23rd percentile for readability.

**Conclusions:** Our findings indicated the GenAI system may generate more highly rated self-management advice than PCPs across most evaluated dimensions but offered no advantage in readability. These findings provide pre-implementation evidence for GenAI as a potential source of structured and adaptive chronic disease self-management support. Patient-level pilot studies and subsequent randomised trials are needed to determine its acceptability and effects on behaviour and clinical outcomes.

## Introduction

Amid global population ageing, chronic non-communicable diseases constitute a leading source of the global disease burden, and their control depends on long-term, continuous self-management in patients’ daily lives^1,2^. Since chronic conditions are long-standing and progressive, self-management for chronic conditions is inherently longitudinal and dynamic according to patients’ clinical status and life circumstances^1^. Plenty of evidence have proved that structured self-management has been shown to improve health behaviors and clinical outcomes and to reduce health-service utilization^1^. Effective self-management, however, demands continuous, personalized support that is inherently resource-intensive, and this nature may render such care inaccessible within primary health care (PHC), where capacity and time are insufficient to deliver adequate chronic disease management support^3,4^.

Since the public release of ChatGPT in late 2022, generative artificial intelligence (GenAI) has been proposed as a potential means of narrowing this gap^3,5^. GenAI, represented prominently by large language model (LLM)–based systems such as ChatGPT, retains the advantages of conventional digital tools, including the capacity to transcend temporal and spatial constraints, while also representing a qualitative advance in technical performance including natural-language interaction, multi-turn dialogue, context-sensitive and individualized content generation, and semantic understanding^6–8^. Together, these capabilities enable it to approximate the communicative behavior of human professionals and to respond flexibly to different patients and evolving problems, and it has been increasingly applied across multiple medical domains including clinical decision support, patient communication and education, clinical documentation, medical training, and biomedical research^7,8^. On this basis, GenAI, in principle, is positioned closer to human professional support than any prior digital tool, with the potential to deliver continuous and dynamically adjusted guidance for chronic disease self-management.

Despite this potential, evidence validating whether GenAI can genuinely support chronic disease self-management, relative to general practice within PHC, remains limited in two principal respects^9,10^. First, existing patient-facing validation studies have largely relied on discrete patient questions or portal messages, with each AI-generated response evaluated independently^11–13^. Although such designs provide evidence regarding response quality in specific encounters, they may fail to capture the continuous nature of real-world chronic self-management, which unfolds over months across multiple stages and requires dynamic adjustment. Second, evaluation of the advice generated by GenAI is dimensionally incomplete, with prior work concentrating on several broad outcomes, such as accuracy, completeness, readability, and empathy^14–16^. Yet few studies have jointly evaluated whether GenAI-generated advice can support behavior change, integrate recommendations across multiple management domains, and adapt as patients’ needs change over time, which are equally important, as they may influence long-term adherence to self-management. Consequently, future research spanning the full course of self-management and employing multidimensional assessment to test the validity of GenAI is warranted.

To address these gaps, we conducted a source-blinded, vignette-based validation study comparing self-management advice generated by a GenAI system with advice written by primary care physicians (PCPs). Standardized vignettes represented three sequential stages of chronic disease self-management including plan initiation, acute adjustment, and long-term maintenance, and advice was assessed using a multidimensional evaluation framework.

## Methods

### Study design

This was a cross-sectional, source-blinded comparative evaluation based on standardized case vignettes, designed to systematically compare the performance of a Gen AI system with that of PCPs in generating self-management advice across the full course of chronic disease management. Reporting adheres to the Chatbot Assessment Reporting Tool (CHART), with the completed checklist provided in the Supplement^17^. Ethical approval was granted by the Peking University Institutional Review Board (IRB00001052-26128).

### Vignette

Vignettes were developed from chronic disease follow-up records routinely collected within the essential public-health service system, from which 15 authentic cases were included. The cases covered the most common chronic conditions in PHC including hypertension and diabetes. To protect privacy, all direct identifiers were removed to preclude re-identification of individual patients by the investigators.

Each vignette comprised baseline patient information and three chronic disease management nodes. The baseline information incorporated multidimensional data, including follow-up records, physical examination, lifestyle assessment, traditional Chinese medicine constitution identification, and ancillary investigations. The scenario evolution across the three nodes was defined in accordance with current clinical guidelines and chronic disease management standards in PHC^18^. The three nodes were configured to evolve sequentially, so as to simulate the continuity and dynamism of real-world self-management: the plan-initiation stage focused on generating a systematic, multidisciplinary self-management plan; the acute-adjustment stage focused on responding to abrupt problems and dynamically modifying the plan during management (e.g., glycaemic or blood-pressure fluctuations, adverse drug reactions, and disruptive life events); and the long-term-maintenance stage focused on declining long-term adherence and psychological maintenance, providing behaviour-maintenance strategies.

To further ensure content validity, all vignettes were independently reviewed by a multidisciplinary expert panel (general practice, endocrinology, and cardiology) for clinical authenticity, representativeness, and node appropriateness before advice generation and scoring. Content validity was quantified using a 4-point scale (1 = not relevant to 4 = highly relevant). Item-level content validity indices (I-CVI) and the scale-level content validity index (S-CVI/Ave) were all ≥ 0.78, indicating good content validity. Detailed results were illustrated in the Table S1.

### Advice generation

For each node of every case, patient-facing self-management advice was generated independently by the PCP group and by the AI system; both received identical vignette text to ensure comparability. A case-level advice set comprised the outputs generated for all three management nodes of a single vignette.

Twenty-two PCPs were recruited from four community health centres in Guangzhou, China. Most participants were women (17/22, 77.3%), held a bachelor’s degree (17/22, 77.3%), and were aged 26–35 years (12/22, 54.5%). Detailed participant characteristics are provided in Table S2. The 15 cases were randomly allocated into four groups (one per centre, four cases each), with case 15 allocated twice. Each physician independently wrote patient-facing self-management advice for the four cases assigned to their centre, under a standardized testing environment in which consultation of guidelines or reference materials and the use of AI tools were prohibited. Responses were entered online through the Wenjuanxing online platform, either by typing or by dictation using a speech-to-text function.

The GenAI system, a large-language-model–based chronic disease management support system whose core model is Qwen3-Max, employs a retrieval-augmented generation (RAG) architecture supported by a domain-specific knowledge base comprising de-identified hospital case data, curated Chinese and international texts on chronic disease management, clinical practice guidelines, expert consensus statements, and other evidence-based resources^19^. The application built around the model was not publicly released as open-source software at the time of the study. For each case, participant data were first structured and summarised, and information relevant to the participant’s current problems, management goals, and safety constraints was retrieved from the knowledge base and supplied to the model as grounding context. Recommendations were then produced through a staged pipeline including safety-risk screening and management-pathway identification, followed by sequential generation of a health-risk assessment, a phased management plan, specific intervention strategies, and daily action recommendations. Each stage was driven by a predefined structured prompt template that specified the model’s role, task objectives, input fields, hierarchy of evidence, target population, output structure, and medical safety boundaries. The model was instructed to generate content exclusively from the information provided and not to fabricate missing data, alter predefined risk classifications, or interpret isolated measurements as diagnostic evidence. All recommendations were generated between April 2026 and May 2026 under a fixed configuration (temperature = 0.7, top-p = 0.9, maximum output = 10,000 tokens), in a single pass and without case-level manual adjustment.

### Evaluation

Advice was assessed using a self-developed seven-dimension framework, with each dimension rated on a 5-point Likert scale (1 = [lowest anchor description], 5 = [highest anchor description]). The dimensions were distributed across the management stages they were designed to assess. Three were evaluated across the full course: (1) medical accuracy—conformity with clinical guidelines and chronic disease management standards in PHC; (2) readability—clarity of language, structural coherence, and suitability for community patients’ comprehension; and (3) behaviour-change potential—provision of clear goals, concrete steps, problem-solving strategies, and actionability. Two were evaluated at the plan-initiation node: (4) individualisation—specific responsiveness to the patient’s condition, lifestyle, and management barriers; and (5) multidisciplinary integration—coordinated guidance across diet, exercise, sleep, medication, and psychological aspects. One was evaluated at the acute-adjustment node: (6) contextual adaptability—fit to the patient’s specific scenario and self-management stage. One was evaluated at the long-term-maintenance node: (7) empathy—respect, understanding, support, and patient-centred communication.

The raters were six experienced clinicians from tertiary Grade-A hospitals—two cardiologists, two endocrinologists, and two general practitioners—with specialty expertise in chronic disease management. Before formal scoring, they completed training and calibration on the operational definitions and scoring anchors of each dimension. Each response was independently scored by three raters, one from each specialty, with no case scored redundantly by two physicians of the same specialty, thereby maintaining a consistent standard within each case. The raters were blinded to the source of each response through uniform preprocessing to standardized plain text by researchers not involved in scoring, and all raters scored independently, without discussion.

### Statistical analysis

The data had a crossed, nested structure: each rating was nested within rater and case, and the two arms of each case (AI system and PCP) constituted paired sources. Independent-sample t tests, which ignore this clustering, were therefore not used, while linear mixed-effects models were applied instead. The primary analysis was restricted to human-expert ratings. For each of the seven dimensions, a separate linear mixed-effects model was fitted with the dimension score as the dependent variable, response source as the fixed effect of interest, and case, rater, complete case-level response set, and PCP as random intercepts, thereby accounting for case difficulty, inter-rater leniency, correlations among ratings of the same response, and repeated responses from the same PCP. We report the estimated marginal means, between-group differences, and 95% confidence intervals. To account for multiple comparisons across the seven dimensions, P values were corrected using the Benjamini–Hochberg false-discovery-rate (FDR) procedure. Inter-rater reliability was assessed using intraclass correlation coefficients (ICC).

Two pre-specified robustness analyses were conducted. First, a case-level paired analysis paired the AI score with the mean PCP score within each case and compared them using paired t and Wilcoxon signed-rank tests. Second, a subgroup positioning analysis computed the percentile rank of the AI mean within the distribution of individual PCP means for each dimension. Analyses were performed in R (version 4.4.1) and in Stata 17.0. All tests were two-sided, and a corrected P < 0.05 was considered statistically significant.

## Results

A total of 103 advice responses including 15 generated by the AI system and 88 by the 22 PCPs were evaluated across the 15 standardized vignettes. Each response was independently scored by three source-blinded experts from tertiary Grade-A hospitals, yielding 309 human-expert ratings, which constituted the dataset for the primary analysis.

The GenAI system performed well across the different stages of chronic disease management, with estimated marginal means generally above 4 on the 1–5 Likert scales, except for readability (Table 1). Across the full course, it scored 4.22 for medical accuracy and 4.18 for behaviour-change potential, but only 2.94 for readability. At the plan-initiation node, it scored 4.12 for individualisation and 4.14 for multidisciplinary integration; at the acute-adjustment node, it scored 4.27 for contextual adaptability; and at the long-term-maintenance node, it scored 4.45 for empathy. In contrast, PCPs were rated in the moderate range, with estimated marginal means ranging from 2.73 for behaviour-change potential to 3.15 for readability.

**Table 1.** Seven-dimension comparison between the AI system and primary care physicians.

| Stage | Dimension | PCP | AI | Difference<br>(95% CI) * | P<br>(FDR) |
|---|---|---|---|---|---|
| Full course | Medical accuracy | 3.06 | 4.22 | 1.16 (0.87, 1.46) | <0.001 |
|  | Readability | 3.15 | 2.94 | -0.22 (-0.54, 0.11) | 0.19 |
|  | Behaviour-change potential | 2.73 | 4.18 | 1.46 (1.11, 1.81) | <0.001 |
| Plan initiation | Individualisation | 2.81 | 4.12 | 1.31 (0.91, 1.71) | <0.001 |
|  | Multidisciplinary integration | 2.79 | 4.14 | 1.35 (1.01, 1.69) | <0.001 |
| Acute adjustment | Contextual adaptability | 2.99 | 4.27 | 1.27 (0.88, 1.66) | <0.001 |
| Long-term maintenance | Empathy | 2.89 | 4.45 | 1.56 (1.15, 1.97) | <0.001 |

\*A positive difference favours the AI system. CI, confidence interval; FDR, Benjamini–Hochberg false-discovery-rate corrected.

In the linear mixed-effects models, the GenAI system scored significantly higher than PCPs on six of the seven dimensions, with all six differences remaining significant after FDR correction (all P<.001). The largest advantage was observed for empathy (adjusted mean difference 1.56, 95% CI 1.15 to 1.97) and behaviour-change potential (1.46, 95% CI 1.11 to 1.81), followed by multidisciplinary integration (1.35, 95% CI 1.01 to 1.69), individualisation (1.31, 95% CI 0.91 to 1.71), contextual adaptability (1.27, 95% CI 0.88 to 1.66), and medical accuracy (1.16, 95% CI 0.87 to 1.46). Readability was the sole exception: the GenAI system scored slightly lower than PCPs, but the difference was not statistically significant (−0.22, 95% CI −0.54 to 0.11; P=.19).

Inter-rater reliability was quantified using two-way random-effects ICCs for absolute agreement. Single-rater reliability ranged from 0.18 to 0.53, and the average-measures reliability for the three-rater scheme ranged from 0.39 to 0.77 (Table 2). Reliability was highest for empathy and behaviour-change potential and lowest for readability, indicating that readability was the most subjectively judged dimension. Systematic rater-leniency differences were further controlled through the rater random effect in the primary models.

**Table 2.** Inter-rater reliability across the seven evaluation dimensions.

| Stage | Dimension | ICC (2,1) | ICC (2,3) |
|---|---|---|---|
| Full course | Medical accuracy | 0.31 | 0.58 |
|  | Readability | 0.18 | 0.39 |
|  | Behaviour-change potential | 0.49 | 0.74 |
| Plan initiation | Individualisation | 0.37 | 0.64 |
|  | Multidisciplinary integration | 0.31 | 0.57 |
| Acute adjustment | Contextual adaptability | 0.35 | 0.61 |
| Long-term maintenance | Empathy | 0.53 | 0.77 |

Note：ICC, intraclass correlation coefficient, estimated from two-way random-effects models for absolute agreement. ICC (2,1) denotes single-rater reliability; ICC (2,3) denotes the average-measures reliability for the three-rater scoring scheme actually used.

Two sensitivity analyses were consistent with the primary results. First, in the case-level paired analysis, the AI system was superior to PCPs on all six content dimensions (paired t and Wilcoxon signed-rank tests, all P < 0.001), whereas readability did not differ significantly (paired t P = 0.32; Wilcoxon P = 0.38), reproducing the primary findings without distributional assumptions (Fig. 1). Second, in the subgroup positioning analysis, the AI mean exceeded the means of all 22 individual PCPs on the six content dimensions (100th percentile), but ranked at approximately the 23rd percentile for readability (i.e., below most PCPs) (Fig. 2). Together, these analyses indicate that the AI advantage is concentrated in content quality and individualised support, whereas its readability does not reach the typical level of a human PCP.

**Figure 1.**
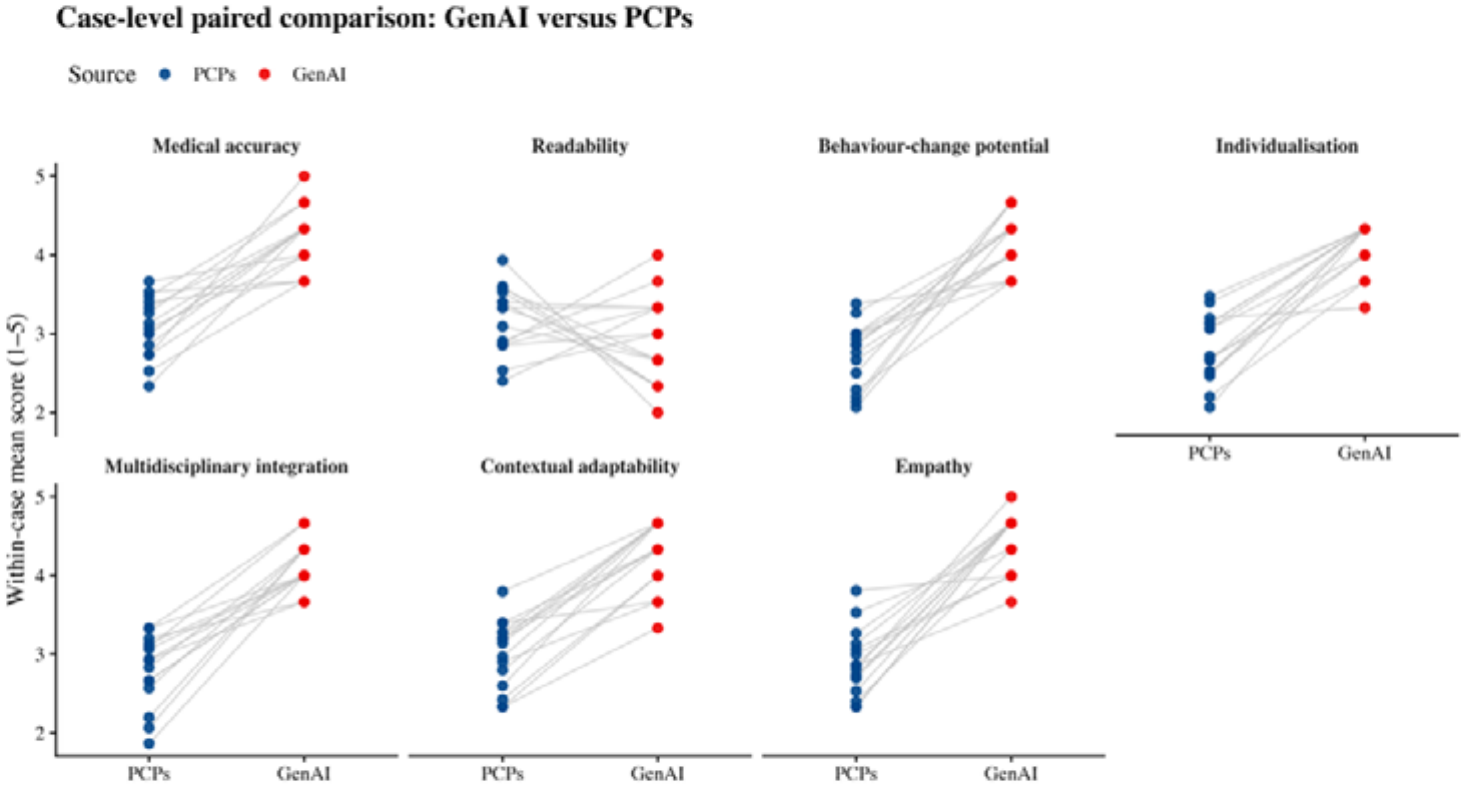
Case-level paired comparison of GenAI and PCP scores

**Figure 2.**
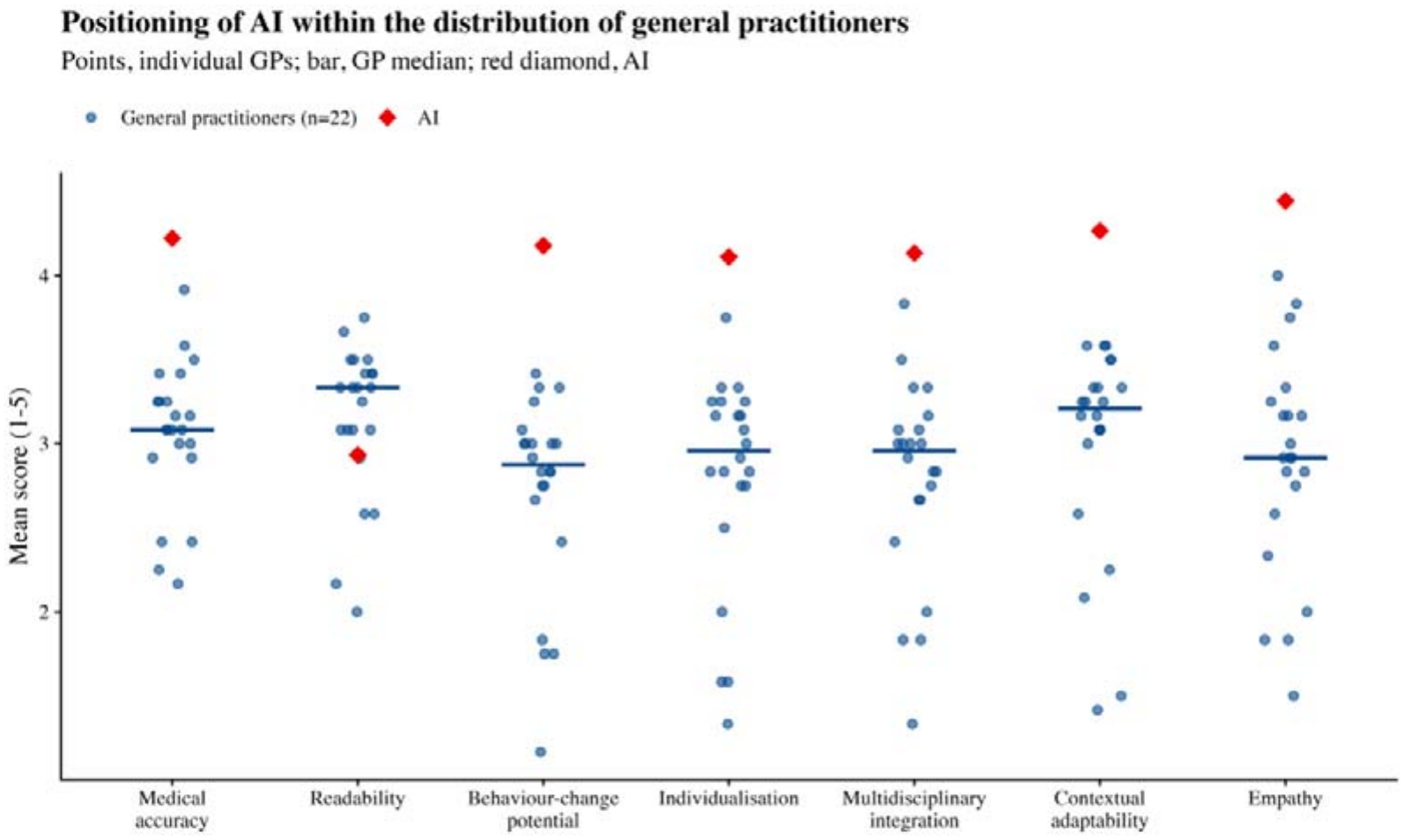
Distribution of the GenAI system within primary care physician scores

## Discussion

GenAI has the potential to address important gaps in chronic disease management by delivering accessible, personalized, and dynamically adjustable support beyond the temporal and resource constraints of routine primary care^8,20^. However, previous validation studies have predominantly focused on isolated, single-turn questions and a limited range of outcomes, particularly medical accuracy and empathy, thereby failing to capture the longitudinal nature and multidimensional requirements of chronic disease self-management^9,10,21^. To address these limitations, we conducted a source-blinded, vignette-based comparative evaluation using sequential clinical scenarios spanning plan initiation, acute adjustment, and long-term maintenance, together with a seven-dimension assessment framework. Across these stages, the GenAI system demonstrated substantially better performance than PCPs in six of the seven evaluated dimensions, whereas no advantage was observed for readability. These findings provide preliminary evidence that the evaluated GenAI system may generate highly rated self-management advice across simulated management stages, supporting further evaluation of its use in patient care.

Beyond confirming previously reported strengths of GenAI in producing accurate and empathetic health communication, our study also extends the evidence to a broader set of capabilities essential for longitudinal chronic disease self-management. Similar to our findings, previous studies have shown that GenAI can provide high-quality, detailed, and supportive responses to patient questions, potentially because large language models can synthesize broad knowledge and generate structured, contextually appropriate, and emotionally supportive language^22–24^. More importantly, it also demonstrated substantial advantages in behavior-change potential, individualization, multidisciplinary integration, and contextual adaptability, dimensions that have received limited attention in prior evaluations, which have largely focused on isolated questions and a narrow range of outcomes^9,10,21^. Such capabilities are especially relevant to chronic disease management, in which patients need to coordinate medication use, diet, physical activity, sleep, psychological well-being, and self-monitoring; adapt their management in response to clinical fluctuations and disruptive life events; and sustain recommended behaviors despite treatment fatigue or declining motivation^1^. These additional capabilities may arise from the ability of GenAI to jointly process clinical indicators, lifestyle characteristics, management barriers, and psychosocial circumstances, and to translate this information into coherent and actionable guidance^8,25,26^. By integrating recommendations across care domains, converting general advice into specific goals and sequenced actions, and adapting support to changing circumstances, GenAI may contribute not merely as a source of health information but as a scalable tool for structured self-management support across different stages of chronic disease management^25,26^.

Readability was the only dimension in which the GenAI system did not outperform PCPs, highlighting a potential trade-off between comprehensive content and cognitive accessibility. Consistent with our finding, an evaluation of real patient in-basket messages found that AI-generated drafts were longer, more lexically diverse, and required a higher reading grade level than responses written by health professionals^13^. However, previous results have varied across clinical contexts: clinicians rated chatbot responses as more readable for cancer-related questions, whereas physician-written responses were preferred for dermatology portal messages^11,15^. These differences may reflect variation in the models, prompts, clinical topics, response lengths, evaluator populations, and definitions of readability used across studies. In our study, the same capacity that enabled the AI system to integrate multiple management domains and generate detailed, individualized recommendations may also have increased information density and cognitive burden. This issue is particularly relevant to chronic disease management because patients need to repeatedly understand and implement recommendations in daily life, often while managing limited health literacy or age-related cognitive challenges. Future patient-facing AI systems may therefore benefit from generating more concise, prioritized, and health-literacy-adaptive advice while retaining essential clinical and behavioral information.

Our findings have two main implications. First, the superior performance of Gen AI across full course of self-management for chronic disease suggests that its role could extend beyond answering isolated health questions. By integrating multiple management domains, translating recommendations into concrete actions, and adapting support across different management stages, GenAI may complement primary care by providing scalable and structured self-management support^27^. Further patient-level studies are needed to determine whether these advantages in expert-rated advice quality translate into sustained behaviour change and improved clinical outcomes. Second, the absence of an advantage in readability indicates that high-quality and comprehensive advice is not necessarily easy for patients to understand and apply. Future implementation may therefore benefit from adapting response length, information density, terminology, and prioritization to patients’ health literacy and cognitive capacity. Combining clinically reliable content with concise and accessible presentation may be important for realizing the potential of Gen AI in long-term chronic disease self-management.

Several limitations must be acknowledged in this study. First, the evaluation was based on standardized case vignettes rather than actual longitudinal patient–AI interactions. Although the three management stages were designed to reflect the evolving nature of chronic disease self-management, they might not fully reproduce the complexity of sustained multi-turn communication, and the study did not assess changes in patient behavior or clinical outcomes^28,29^. Moreover, the ratings were primarily provided by health professionals, whose assessments may not fully represent patients’ experiences. Patient-level evaluations are needed to assess the feasibility and acceptability of the generated advice. Second, the study included only 15 cases involving diabetes and hypertension within the context of PHC. The findings may not generalize to other chronic diseases or patients with complex multimorbidity. Future evaluations using larger and more diverse case samples would help establish the external validity of the findings. Third, the results were derived from a single AI system under a specific prompting and generation configuration. Since performance may vary across models, prompts, knowledge sources, and generation parameters, the findings should be interpreted as evidence for the evaluated system rather than for GenAI in general^30^. External validation across different models and repeated evaluation following major system updates may be needed to determine the robustness and temporal stability of its performance.

## Conclusion

In this source-blinded, vignette-based comparative evaluation, the GenAI system outperformed PCPs in six of seven dimensions across simulated stages of chronic disease self-management, while showing no advantage in readability. These findings provide pre-implementation evidence that GenAI may offer scalable, structured, and adaptive support extending beyond the provision of health information, although its content needs to be made more accessible to patients with varying levels of health literacy. Patient-level pilot evaluation and subsequent randomized controlled trials are required to determine whether these advantages in advice quality translate into acceptable use, sustained behavior change, and improved clinical outcomes.

## Supporting information

checklist

## Data availability

The de-identified data supporting the findings of this study are available from the corresponding authors upon reasonable request, subject to applicable ethical and institutional requirements. The original patient records are not publicly available to protect patient privacy.

## Code availability

The code used for data processing and statistical analysis is available from the corresponding authors upon reasonable request.

## Ethical approval

This study was approved by the Peking University Institutional Review Board (approval number: IRB00001052-26128).

## Author contributions

Z.Z., W.C., and X.M. conceived and designed the study and coordinated physician recruitment and data collection. Z.Z. developed the standardized vignettes and coordinated the content-validity assessment. H.Y., and Q.W. developed and configured the GenAI system and generated the AI responses. Z.Z. developed the statistical analysis plan, analysed the data, and prepared the tables and figures. Z.Z. drafted the manuscript. W.C. and X.M. supervised the study. All authors contributed to the interpretation of the findings, critically revised the manuscript, and approved the final version. X.M. is the guarantor of the study and takes responsibility for the integrity of the work as a whole.

