## supplement for "Generative AI Versus Primary Care Physicians from Plan Initiation to Long-Term Maintenance of Chronic Disease Self-Management: A Vignette-Based Validation Study"

Supplementary Table S1. Content validity indices for the 15 standardized case vignettes

| Vignette | Clinical authenticity, I-CVI | Representativeness, I-CVI | Stage appropriateness,  I-CVI | Mean across dimensions |
| --- | --- | --- | --- | --- |
| 1 | 1.000 | 1.000 | 1.000 | 1.000 |
| 2 | 1.000 | 1.000 | 1.000 | 1.000 |
| 3 | 1.000 | 1.000 | 1.000 | 1.000 |
| 4 | 1.000 | 1.000 | 1.000 | 1.000 |
| 5 | 0.667 | 1.000 | 1.000 | 0.889 |
| 6 | 1.000 | 1.000 | 1.000 | 1.000 |
| 7 | 0.667 | 0.667 | 1.000 | 0.778 |
| 8 | 1.000 | 1.000 | 1.000 | 1.000 |
| 9 | 1.000 | 1.000 | 1.000 | 1.000 |
| 10 | 1.000 | 1.000 | 1.000 | 1.000 |
| 11 | 1.000 | 1.000 | 1.000 | 1.000 |
| 12 | 1.000 | 1.000 | 0.667 | 0.889 |
| 13 | 1.000 | 0.667 | 0.667 | 0.778 |
| 14 | 1.000 | 1.000 | 1.000 | 1.000 |
| 15 | 1.000 | 1.000 | 1.000 | 1.000 |
| S-CVI/Ave | 0.956 | 0.956 | 0.956 | 0.956 |

*Note:* I-CVI, item-level content validity index; S-CVI/Ave, scale-level content validity index calculated using the averaging method. Each vignette was independently assessed by three experts, one each from general practice, endocrinology, and cardiology, using a four-point scale. Ratings of 3 or 4 indicated endorsement.

Supplementary Table S2. Characteristics of participating primary care physicians

| Characteristic | PCPs (N = 22) |
| --- | --- |
| Sex, n (%) |  |
| Female | 17 (77.3) |
| Male | 5 (22.7) |
| Age, years, n (%) |  |
| 26–35 | 12 (54.5) |
| 36–45 | 7 (31.8) |
| 46–55 | 3 (13.6) |
| Educational attainment, n (%) |  |
| Bachelor’s degree | 17 (77.3) |
| Master’s degree | 5 (22.7) |
| Professional title, n (%) |  |
| Junior | 7 (31.8) |
| Intermediate | 12 (54.5) |
| Associate senior | 3 (13.6) |
| Years in practice, n (%) |  |
| ≤5 | 13 (59.1) |
| 6–10 | 5 (22.7) |
| 11–20 | 2 (9.1) |
| >20 | 2 (9.1) |
| Number of contracted patients, n (%) |  |
| <500 | 4 (18.2) |
| 500–1,000 | 2 (9.1) |
| 1,001–1,500 | 9 (40.9) |
| >1,500 | 7 (31.8) |
| Recorded task-completion time, min |  |
| Median (IQR) | 94.8 (48.0–137.3) |
| Range | 4.1–254.8 |

*Note:* Data are presented as n (%) unless otherwise indicated. IQR, interquartile range; PCP, primary care physician.
