## Supplementary material for "Generative AI Versus Primary Care Physicians from Plan Initiation to Long-Term Maintenance of Chronic Disease Self-Management: A Vignette-Based Validation Study": checklist

### CHART: Chatbot Assessment Reporting Tool

Reporting Checklist for Chatbot Health Advice Studies

| Heading | Item # | Item | Page # |
|---|---|---|---|
| **Title & Abstract** | | | |
| Title | 1a | State that the study is assessing one or more generative AI-driven chatbots for clinical evidence or health advice. | 1 |
| Abstract/Summary | 1b | Apply a structured format, if applicable. | 2 |
| **Introduction** | | | |
| Background | 2a | State the scientific background, rationale, and healthcare context for evaluating the generative AI-driven chatbot(s), referencing relevant literature when applicable. | 3-4 |
| | 2b | State the aims and research questions including the target audience, intervention, comparator(s), and outcome(s). | 4-7 |
| **Methods** | | | |
| Model Identifiers | 3a | State the name and version identifier(s) of the generative AI model(s) and chatbot(s) under evaluation, as well as their date of release or last update. | 6 |
| | 3b | State whether the generative AI model(s) and chatbot(s) are open-source or closed-source/proprietary. | 6 |
| Model Details | 4a | State whether the generative AI model was a base model or a novel base model, tuned model, or fine-tuned model. | 6 |
| | 4b | If a base model is used, cite its development in sufficient detail to identify the model. | 6 |
| | 4c | If a novel base model, tuned model, or fine-tuned model is used, describe the pre- and/or postimplementation/deployment data and parameters. | 6 |
| Prompt Engineering | 5a | Describe the evolution of study prompt development. | 6 |
| | 5ai | Describe the sources of prompts. | 6 |
| | 5aii | State the number and characteristics of the individual(s) involved in prompt engineering. | 6 |
| | 5aiii | Provide details of any patient and public involvement during prompt engineering. | NA |
| | 5b | Provide study prompts. | 6 |
| Query Strategy | 6a | State route of access to generative AI model. | 6 |
| | 6b | State the date(s) and location(s) of queries for the generative AI-driven chatbot(s) including the day, month, and year as well as city and country. | 6 |
| | 6c | Describe whether prompts were input into separate chat session(s). | 6 |
| | 6d | Provide all generative AI-driven chatbot output/responses. | 17 |
| Performance Evaluation | 7a | Define the ground truth or reference standard used to define successful generative AI-driven chatbot performance. | 6-7 |
| | 7b | Describe the process undertaken for generative AI-driven chatbot performance evaluation. | 6-7 |
| | 7bi | State the number and characteristics of team members involved in performance evaluation. | 7 |
| | 7bii | Provide details of any patients and public | NA |

| Heading | Item # | Item | Page # |
|---|---|---|---|
| | | involvement during the evaluation process. | |
| | 7biii | State whether evaluators were blinded to the identity of the generative AI-driven chatbot(s) under assessment. | 7 |
| Sample Size | 8 | Report how the sample size was determined. | NA |
| Data Analysis | 9a | Describe statistical analysis methods, including any evaluation of reproducibility of generative AI-driven chatbot responses. | 7-8 |
| | 9ai | Report the measures used for performance evaluation. | 6-8 |
| **Results** | | | |
| | 10a | Report the performance evaluation undertaken including the alignment between generative AI-driven chatbot output and ground truth or reference standard using quantitative or mixed methods approaches as applicable. | 8-9 |
| | 10b | For responses deviating from the ground truth or reference standard, state the nature of the difference(s). | 8-9 |
| | 10c | Report the evaluation for potentially harmful, biased, or misleading responses. | 8-9 |
| **Discussion** | | | |
| | 11a | Interpret study findings in the context of relevant evidence. | 9-11 |
| | 11b | Describe the strengths and limitations of the study. | 11-12 |
| | 11c | Describe the potential implications for practice, education, policy, regulation, and research. | 11-12 |
| **Open Science** | | | |
| Disclosures | 12a | Report any relevant conflicts of interest for all authors. | 17 |
| Funding | 12b | Report sources of funding and their role in the conduct and reporting of the study. | 17 |
| Ethics | 12c | Describe the process undertaken for ethical approval. | 17 |
| | 12ci | Describe the measures taken to safeguard data privacy of patient health information, as applicable. | 17 |
| | 12cii | State whether permission/licensing was obtained for the use of original, copyrighted data. | NA |
| Protocol | 12d | Provide a study protocol. | NA |
| Data availability | 12e | State where study data, code repository, and model parameters can be accessed. | 17 |

**If in supplementary appendix, indicate "supp" and appendix #, if applicable.*
